# The Ophthalmic Surgical Backlog caused by the COVID-19 Pandemic: A population-based and microsimulation modelling study

**DOI:** 10.1101/2021.03.25.21254375

**Authors:** Tina Felfeli, Raphael Ximenes, David MJ Naimark, Philip L Hooper, Robert J Campbell, Sherif R El-Defrawy, Beate Sander

## Abstract

**Background:** Jurisdictions worldwide ramped down ophthalmic surgeries to mitigate the effects of coronavirus disease 2019 (COVID-19), creating a global surgical backlog. We sought to predict the long-term impact of COVID-19 on ophthalmology surgical care delivery.

**Methods:** This is a population-based and a microsimulation modelling study. Provincial administrative data from January 2019 to May 2021 was used to estimate the backlog size and wait-times following the COVID-19 pandemic. For the post-pandemic recovery phase, we estimated the resources required to clear the backlog of patients accumulated on the waitlist during the pandemic.

**Results:** A total of 56,923 patients were on the waitlist in the province of Ontario awaiting non-emergency ophthalmic surgery as of March 15, 2020. The number of non-emergency surgeries performed in the province decreased by 45-98% from March to May 2020, and 48-80% from April to May 2021 compared to the same months in 2019. By 2 years and 3 years, the overall estimated number of patients awaiting surgery grew by 129% and 150%, respectively. The estimated mean wait-time for patients for all subspecialty surgeries increased to 282 (SD 91) in March 2023 compared to 94 days (SD 97) in 2019. The provincial monthly additional resources required to clear the backlog by March 2023 was estimated to be a 34% escalation from the pre-pandemic volumes (4,626 additional surgeries).

**Interpretation:** The magnitude of the ophthalmic surgical backlog from COVID-19 has important implications for the recovery phase. The estimates from this microsimulation modelling can be adapted to other jurisdictions to assist with recovery planning for vision saving surgeries.

## Introduction

The global spread of coronavirus disease 2019 (COVID-19) has led to major disruption of elective or non-urgent surgical procedures globally.^1–4^ Across 190 countries, it is estimated that more than 28 million surgeries were postponed in the immediate months following the COVID-19 shutdowns.^3^ Starting March 15, 2020 in Ontario, Canadian hospitals began ramping down the number of scheduled surgeries and procedures, including ophthalmic surgeries.^5^ Large backlogs of patients accrued as a consequence of the lockdown and may lead to deteriorating quality of life as well as development of irreversible vision impairment. Most recently, the increase in turnover times arising from additional donning and doffing of personal protective equipment and cleaning protocols for COVID-19 has also decreased the number of surgeries performed daily.^6^ With the recommencement of elective activities, patients are likely to be prioritised by clinical urgency, which may further lengthen delays for patients with non-imminently vision-threatening but progressive eye conditions. For example, delays in care have been shown to have unintended consequences for outcomes of retinal detachments^7^ and neovascular age-related macular degeneration.^8^

Predicting the long-term backlog created for ophthalmic surgeries as a result of the COVID-19 pandemic will provide guidance for health care systems to adequately prepare for the ongoing pandemic and post-pandemic recovery phase. Microsimulation modelling offers the ability to track individual patients as they traverse the hospital system and run scenarios to assess the impact of the reductions in ophthalmic surgeries on wait times and consequences of delayed surgeries while taking into account the interactions between prioritization of surgeries performed based on urgency, specialty availability of resources and incidence case growth.^9–11^ Health system decision-analytic models, can forecast health care utilization, patient outcomes over time, dynamic resource constraints, queuing and priority setting, and the consequences of resource constraints, which are useful in answering future-minded health system questions.^10^ With the implementation of the provincial Wait Time Strategy database from Ontario Health, all wait times for patients are comprehensively captured for the Ontario population of 14.7 million.^12^ The Wait Time Information System database provides a unique opportunity to study the impact of the COVID-19 pandemic on the wait-time for various subspecialty surgeries including vitreoretinal, glaucoma, cornea, cataract, oculoplastics, and strabismus surgery.^13^

Ophthalmology is a high-volume specialty with already large waitlists of patients prior to the pandemic. With the recommencement of elective activities, patients are likely to be prioritized by clinical urgency, which may further lengthen delays for patients with non-imminently vision-threatening but progressive eye conditions. It is important to understand the impact of interrupting surgeries on the waiting time for ophthalmic surgeries. Given the patient hesitancy and delays in routine follow-ups for eye care,^14^ the accurate estimates of patients waiting for non-emergency ophthalmic surgery is not available in Ontario, as such, prediction models informed by historical data are the optimal means of determining the effect of the COVID-19 pandemic on wait lists. Herein, we present the first ophthalmology microsimulation prediction model informed by province-wide databases, which aims to project the long-term impact of COVID-19 on ophthalmology surgical volumes, wait times and post-pandemic recovery phase in Ontario, Canada. A better understanding of the magnitude of the surgical backlog from COVID-19 will serve as a framework for modelling surgical backlog recovery planning for vision saving surgeries in other jurisdictions.

## Methods

An individual-level, discrete-time, microsimulation model was developed to simulate adult patients (18 years and older) waiting for ophthalmic surgery at the start of the COVID-19 pandemic. The primary outcomes of the study were the number of patients awaiting non-emergency ophthalmic surgery per month and the time to surgery (number of days on the wait list) based on the subspecialty surgery type and level of urgency. In addition, the escalation in resources required to clear the backlog during the post-pandemic recovery phase was estimated. Waiver of ethics review for use of de-identified population-level data for the model was permitted by the University of Toronto Institutional Review Board for Human Subjects Research and the study procedures adhered to the tenets of the Declaration of Helsinki.

### Data Sources

Provincial administrative data from January 1, 2019 to May 30, 2021 was used from the Province of Ontario Wait Time Information System database to parameterize the model. Ontario Health (Cancer Care Ontario) is authorized to collect population-level data for the purpose of monitoring allocation of resources and delivery of services. The database captures patients from five health regions (West, Central, Toronto, East and North)^14^ who are on the wait list queue for non-emergency surgical procedures as of the first day of each month as well as the number of new cases added to the wait list and number of completed surgeries performed. Additionally, the database captures the wait time in days (mean and standard deviation) for patients on the wait list. All surgical cases in Ontario are standardized into priority levels 1 to 4 (with 1 being the most urgent and irreversible causes of vision loss) with associated maximum surgical wait time targets. These priority level definitions for wait times reflect the need to accelerate care that minimizes impact of disability on patients and are accepted by the federal, provincial, and territorial ministers of health (<u>Supplemental Table 1</u>).^12^ Given the urgency of Priority 1 cases, they are not added to a waitlist and thus not captured in the Ontario Wait Time Information System database.

Facility-level data from March 1, 2018 to August 1, 2020 and May 30, 2021 for four Toronto Central Local Health Integration Network academic hospitals (Kensington Vision and Research Centre, Mount Sinai Hospital, Sunnybrook Health Sciences Centre and Toronto Western Hospital) was used to capture the details on number of patients and wait times for emergency cases (Priority 1) undergoing surgery. These centres consist of hospital-based and stand-alone centres which represent the variety of ophthalmic surgical centres across Ontario. Lastly, guidelines from the quality-based procedures for subspecialty surgery from the Ministry of Health and Long-Term Care^15^ and The Vision Task Force, the literature and expert opinion from ophthalmology specialists were consulted to determine an order of priority based on urgency for vitreoretinal surgery,^16^ glaucoma, cornea,^17^ cataract surgery,^15^ oculoplastics and adult strabismus surgery. For example, urgent vitreoretinal surgical cases were given a higher priority than urgent oculoplastics cases with an acceptable wait time of less than a week prior to deterioration based on the current literature on decline in functional outcomes of macula sparing and involving retinal detachments.^18–21^ A summary of the parameters used in the model for each of the subspecialties and urgency levels is outlined in <u>Supplemental Table 2</u>.

### Model Structure

The time-steps of the microsimulation model were each one day long. The subspecialties included in the model were cataract surgery (cataract and combination cataract and other procedures), retina (vitrectomy and other vitreoretinal surgery), glaucoma surgery (glaucoma filter/seton and other glaucoma surgeries), corneal surgery (corneal transplant and other cornea surgery), oculoplastics and adult strabismus surgery, each based on their unique characteristics. On initiation of simulations, individuals representing the existing waitlist as of March 15, 2020 based on real data entered the model. On each subsequent day, estimates of new urgent and semi-urgent/non-urgent cases informed by real data from 2019-2020 entered the model and were added to the surgical waitlist (Figure 1).

**Figure 1.**
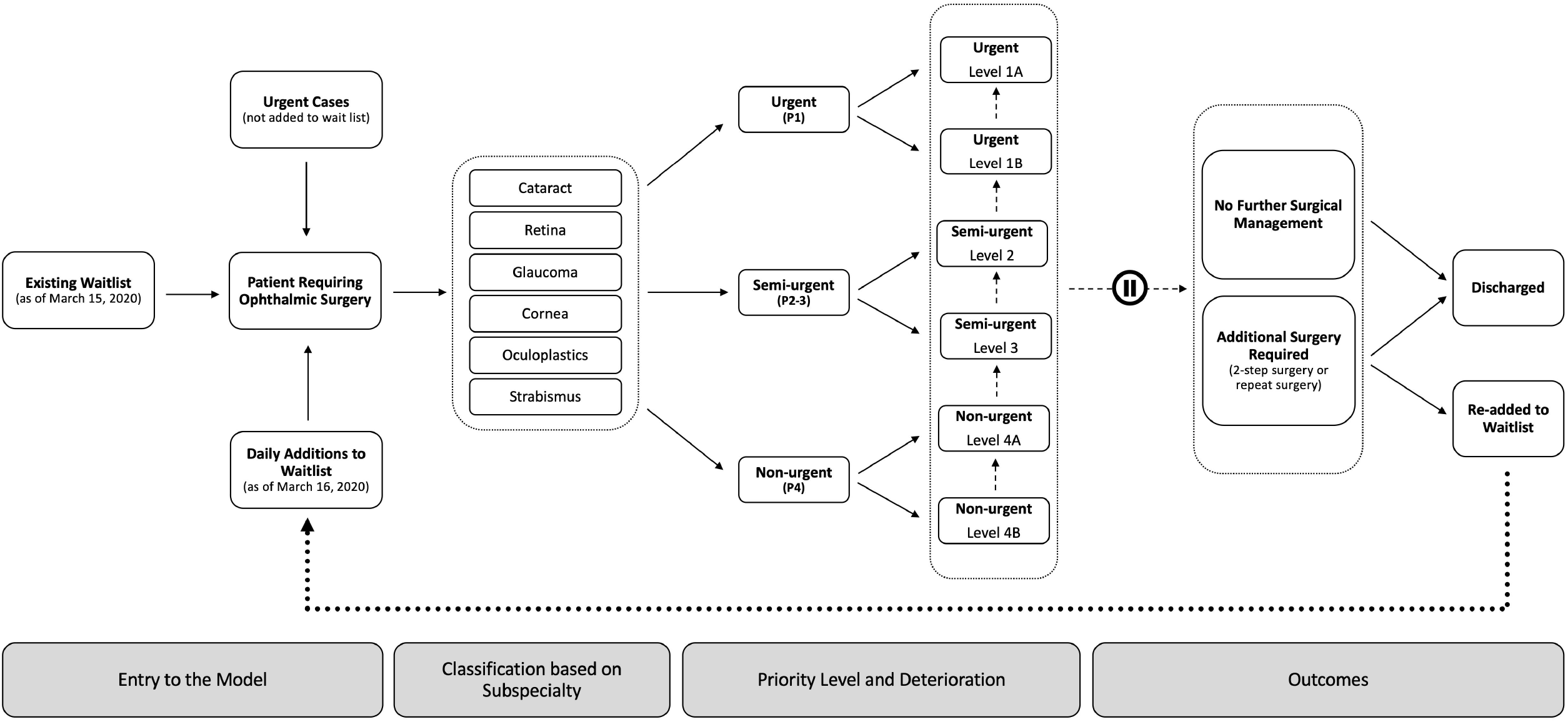
Model schematic depicting patient flow for cases requiring subspecialty ophthalmic surgery. Two entry streams for patients including urgent cases and surgical wait list (consists of existing waitlist prior to the pandemic and daily referrals following declaration of the pandemic). The stop nodes (=) represent resource constraint for ophthalmic subspecialty surgery. For patients in semi-urgent and non-urgent classifications, there is a deterioration and increase in urgency priority (as indicated by the dashed line) for surgery as the maximum wait time is reached (highest priority given to ‘Level 1’). This was done to account for the risk of vision impairment associated with delays in surgical repair. Patients move to the ‘Outcomes’ health states following surgery only when resources become available. Those requiring additional surgical interventions will re-enter the model (as indicated by the dotted line).

On each day, a fixed number of procedures were scheduled informed by the number of procedures available per month in the Province based on estimates from real data. The surgeries occurred seven days of the week to account for urgent surgeries after hours and on weekends. Patients with the highest urgency (Priority 1) underwent subspecialty surgery, followed by semi-urgent groups (Priority 2 and 3) and then non-urgent (Priority Level 4) if the necessary resources were available. Within each urgency level (the urgent, semi-urgent and non-urgent classifications), the prioritization for allocation of surgery was further broken down to multiple levels (1A, 1B, 2, 3, 4A and 4B) based on length of wait time. Patients remained on the waitlist until the next available resource for surgery became available. Following each surgical intervention, patients were assigned a probability for full recovery with ‘No Further Surgical Management’ required. The remaining patients in the ‘Additional Surgery Required’ category underwent pre-specified 2-step surgery (e.g., silicone oil removal and intraocular lens insertion for patients left aphakic following initial surgery) or repeat surgery for those with a suboptimal surgical outcome following the initial surgery (e.g., intraocular lens repositioning). Similar to real life, it was assumed that each person could have a maximum three surgeries, as repeating more than three surgeries for the same condition is extremely rare. The model assumed that all surgeries performed were unilateral. Patients moved up through the urgency prioritization levels based on time on the wait list to account for deterioration of vision status with long delays in surgical repair.

### Backlog Trajectory and Recovery Plan

Simulations started on March 15, 2020 with an end date on March 1, 2023. Outcomes were accrued over a time horizon of 3 years (35.5 months). For the pandemic phase (base case) representing the backlog created as a result of the COVID-19 pandemic shutdowns in the province (first [March 2020] and third [April 2021] waves), the number of available resources for January 2019 to May 2021 from the Wait Time Information System database was used. In order to reflect hospital resource availabilities for ophthalmic surgery, the model was set up with specific constraints (set number of surgeries/operating room available). The number of new patients awaiting surgery was calculated based on historical numbers from 2019 in the Wait Time Information System database up to November 1, 2020. Following November 1, 2020, the number of new patients added to the wait list and the surgical resources was based the disease incidence, previous annual growth of waitlist and Government of Ontario population projections (1.3% increase 2021 and 1.4% increase 2022).^22^

For the post-pandemic recovery phase, we estimated the resources required to clear the backlog of patients accumulated on the waitlist since the pandemic on March 15, 2020 for different time horizons.

### Model Validation

The estimates of wait time obtained from the model for March to November 2019 were compared with the historical data from Wait Time Information System database for the same time period. This was done to ensure that the model could adequately predict future wait times for patients awaiting surgery. A comparison of the projected wait times from the model for the months of March to November 2019 demonstrated no significant differences in the findings between the model and historical data from the Wait Time Information System database (124.40 [SD 70.77] vs. 122.28 [134.61]). These findings support the accuracy of the model in projecting the backlog as a result of the pandemic for 2021-2023.

### Analysis and Sensitivity Analysis

To account for the variability and uncertainty in the inputs, a probabilistic sensitivity analysis (PSA) was conducted. The PSA was run 50 times for each scenario model with approximately 240,000 patients over each year. The PSA incorporates variability in the parameters (as a part of the probabilistic sensitivity analysis) at two levels: 1) patient characteristics, and 2) surgical parameters. Individual-level models incorporate stochasticity, or randomness which reflects uncertainty from random processes. Each of the 50 scenarios run by the model produce a different result, but over a large number of simulations, the results should converge to the average result from a deterministic model. The robustness of the findings despite stochasticity of the parameters is depicted by the clustering of the various scenarios, which suggests that variations in various parameters do not alter the overall findings of the model. The outcomes for all trials were calculated as a mean and standard deviation (SD). All modelling and analyses were conducted using TreeAge Pro 2021 (TreeAge Software, Williamstown, Mass). The mean output from the microsimulation was visualized using the R Statistical Program (version 4.0.4).

## Results

As of March 15, 2020, there were 56,923 patients on the waitlist in the province of Ontario awaiting non-emergency ophthalmic surgery. On average, the monthly number of non-emergency cases for January 2019-February 2020 added to the wait list and undergoing surgery were 14,176 and 13,659, respectively. A summary of the monthly wait list queue, surgical throughputs, newly added cases, and wait times for ophthalmic surgery in Ontario for January 2019-February 2020 are presented in Table 1. The number of non-emergency surgeries performed in the province decreased by 45%, 98% and 97% in March, April and May 2020 (first wave), respectively, compared to the same months in 2019. The number of non-emergency surgeries performed in the province decreased by 48% and 80% in April and May 2021 (third wave), respectively, compared to the same months in 2019. Figure 2 depicts the comparison of the surgical throughput during the pandemic phase compared to historical data from 2019.

**Table 1.** Monthly real data wait list queue, surgical throughputs, newly added cases, and wait times for ophthalmic surgery in Ontario from January 2019 to February 2020 based on subspecialty and priority level. Data source: Wait Times Information System, Ontario Health (Cancer Care Ontario). P2–P4 indicates priority level 2 to 4.

|  | All Subspecialties | Cataract | Retina | Glaucoma | Cornea | Oculoplastics | Strabismus |
| --- | --- | --- | --- | --- | --- | --- | --- |
| Number of Patients Waiting for Surgery – No. |  |  |  |  |  |  |  |
| All Urgencies<sup>a</sup> | 58,380 | 54,030 | 1,154 | 592 | 833 | 1,247 | 524 |
| Semi-urgent (P2-3) | 4,688 | 3,983 | 364 | 190 | 45 | 66 | 38 |
| Non-urgent (P4) | 53,692 | 50,047 | 789 | 402 | 788 | 1,181 | 486 |
| Wait Time, days – mean (SD) |  |  |  |  |  |  |  |
| All Urgencies<sup>a</sup> | 89.58 (95.94) | 93.07 (97.5) | 37.79 (58.87) | 52.01 (60.92) | 92.71 (93.54) | 73.24 (65.85) | 109.34 (101.28) |
| Semi-urgent (P2-3) | 62.77 (92.81) | 80.61 (103.20) | 23.64 (46.04) | 30.63 (35.77) | 37.6 (43.76) | 31.19 (33.6) | 84.59 (145.44) |
| Non-urgent (P4) | 92.64 (95.79) | 94.09 (96.94) | 51.59 (64.33) | 67.51 (70.29) | 98.6 (95.78) | 79.86 (67.22) | 109.1 (88.48) |
| Cases Added per Month – No. |  |  |  |  |  |  |  |
| All Urgencies<sup>a</sup> | 14,176 | 12,697 | 671 | 230 | 179 | 279 | 115 |
| Semi-urgent (P2-3) | 1,657 | 1,155 | 330 | 100 | 24 | 36 | 9 |
| Non-urgent (P4) | 12,520 | 11,542 | 341 | 130 | 155 | 243 | 106 |
| Cases Completed per Month – No. |  |  |  |  |  |  |  |
| All Urgencies<sup>a</sup> | 13,661 | 12,271 | 631 | 220 | 170 | 254 | 109 |
| Semi-urgent (P2-3) | 1,390 | 923 | 312 | 91 | 17 | 34 | 9 |
| Non-urgent (P4) | 12,271 | 11,348 | 319 | 129 | 153 | 220 | 100 |

<sup>a</sup>All urgencies includes semi-urgent and non-urgent cases. Urgent cases are not included as they are not adequately documented through Cancer Care Ontario. No., Number.

**Figure 2.**
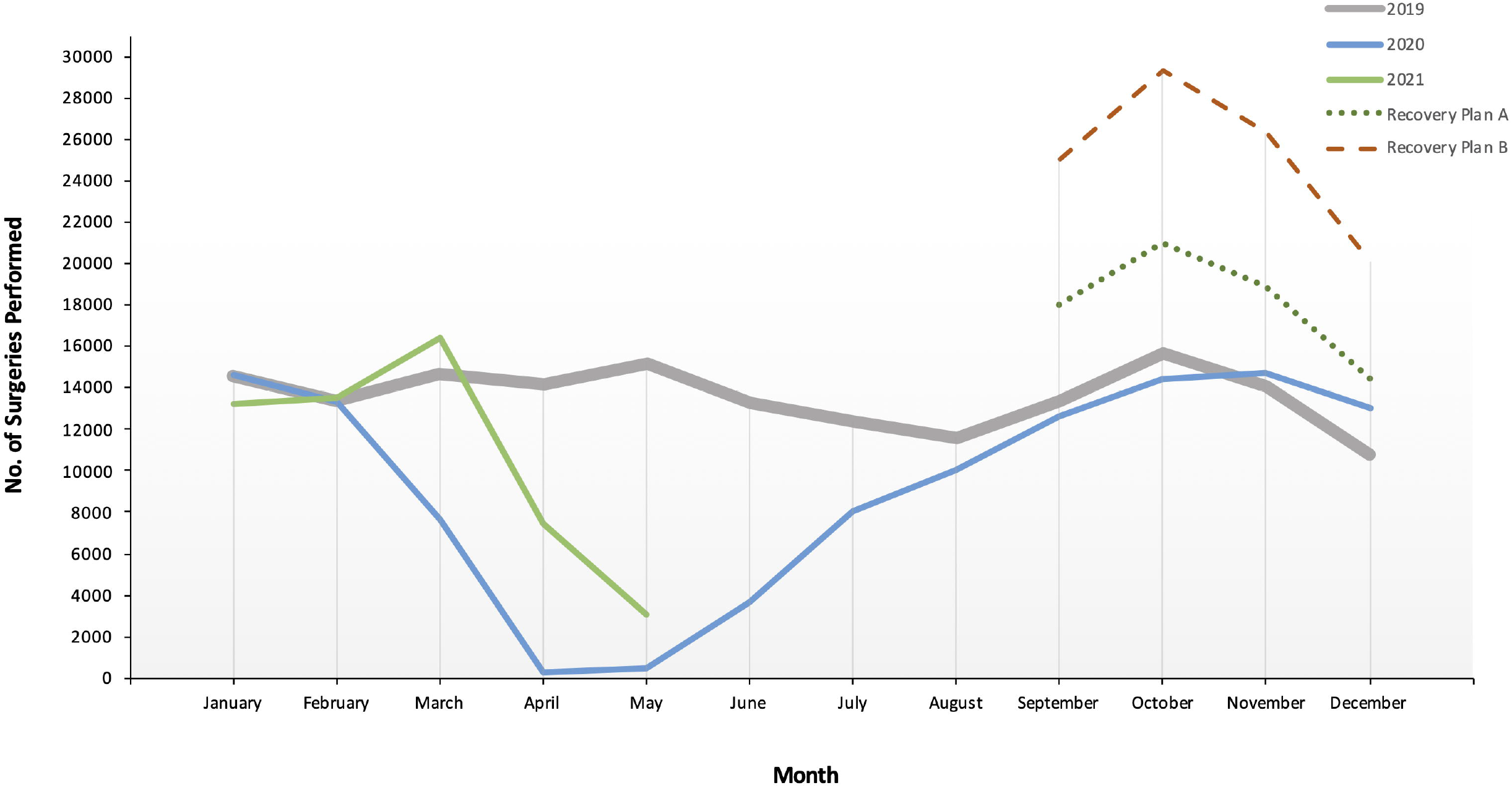
Monthly surgical throughputs based on real data following the pandemic (solid blue line=2020; solid green line=2021) compared to 2019 (solid grey line). The dotted line demonstrates the model-estimated monthly increase in surgical resources required to clear the backlog created as a result of COVID-19 over a two-year period starting in September 2021 (Recovery Plan A). The dashed line demonstrates the monthly increase in surgical resources required to clear the backlog over a one-year period starting in September 2021 (Recovery Plan B). These Recovery Plan results demonstrate the degree of escalation in resource availability required to return to the pre-pandemic wait list queue and wait times for ophthalmic surgery. Note that only the first months of the Recovery Plans are depicted in the graph.

### Model Projections

The total number of patients awaiting surgery at exactly 1 year following the pandemic increased by 112% (62,503 additional cases) in February 2021 compared to February 2020. By 2 years, the overall number of patients awaiting surgery grew by 129% from February 2020. More specifically, the number of patients awaiting cataract, vitreoretinal, glaucoma, cornea, oculoplastics and strabismus surgeries grew by 140%, 133%, 171%, 175%, 115% and 340%, respectively at 2 years following the pandemic. By 3 years, the overall number of patients awaiting surgery grew by 150% from February 2020. Provincial estimates of the backlog size by surgical subspecialty type over 3 years following the pandemic are presented in Figure 3. Overall, the growth in the backlog as a result of the number of patients awaiting surgery was driven by the volume of non-urgent cases (<u>Supplemental Figure 1</u>).

**Figure 3.**
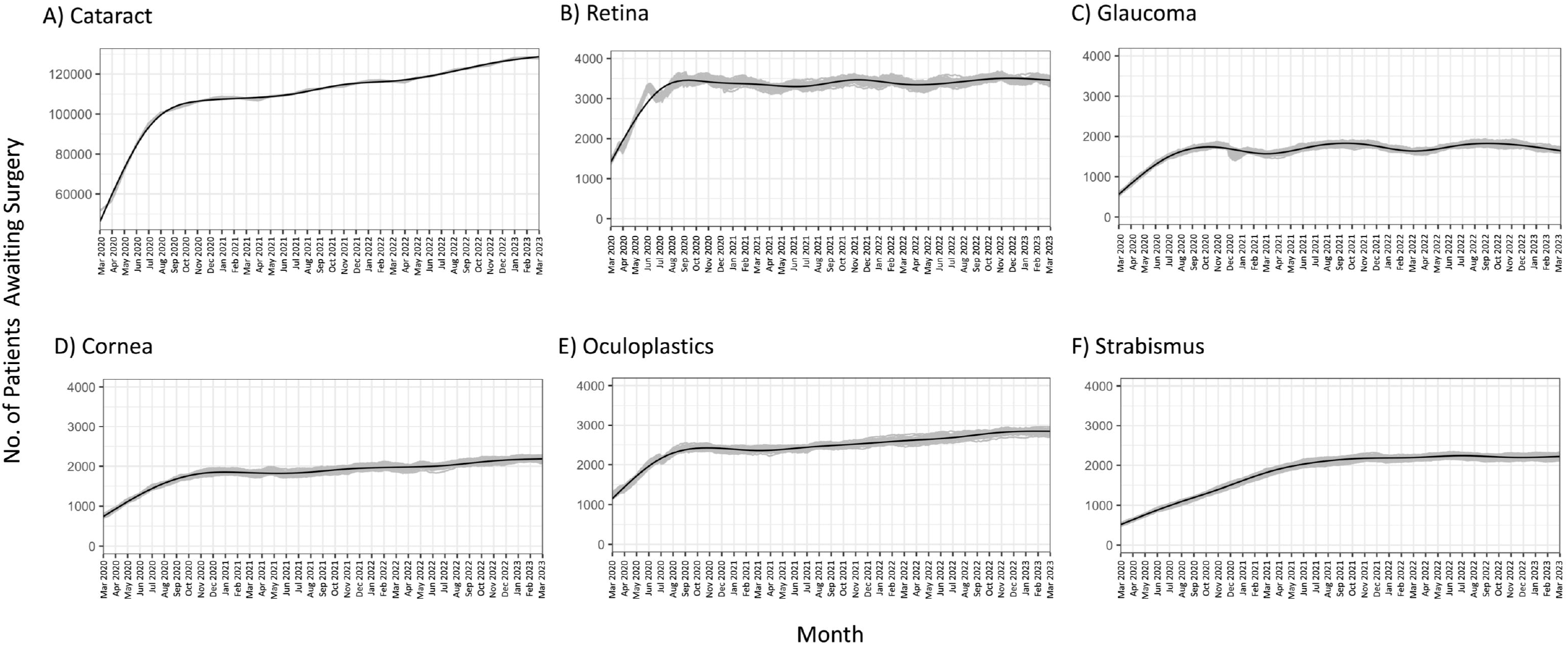
Monthly model-estimated accumulation of patients awaiting surgery for all surgeries and subspecialty types including cataract surgery (A, cataract and combination cataract and other procedures), retina surgery (B, vitrectomy and other vitreoretinal surgery), glaucoma surgery (C, glaucoma filter/seton and other glaucoma surgeries), corneal surgery (D, corneal transplant and other cornea surgery), oculoplastics (E) and adult strabismus surgery (F) since March 2020, to March 2023. The simulations were run 50 times (variations in projected estimated represented by grey lines) for a total of 240,000 patients. Note that the y-axis scale for cataract surgery is different than other subspecialty groups. No., number.

The mean wait time for patients for all subspecialty surgeries increased to 282 (SD 91) in March 2023 compared to 94.41 days (SD 97.42) in 2019 (Table 2). The estimated time to surgery for the initial patients on the wait list at the start of the pandemic was 197.28 (SD 95.09) days. Of the 56,047 patients on the waitlist for semi-urgent and non-urgent surgery at the start of the pandemic, the results suggested that 99% had surgery within 12 months (<u>Supplemental Figure 2</u>).

**Table 2.**
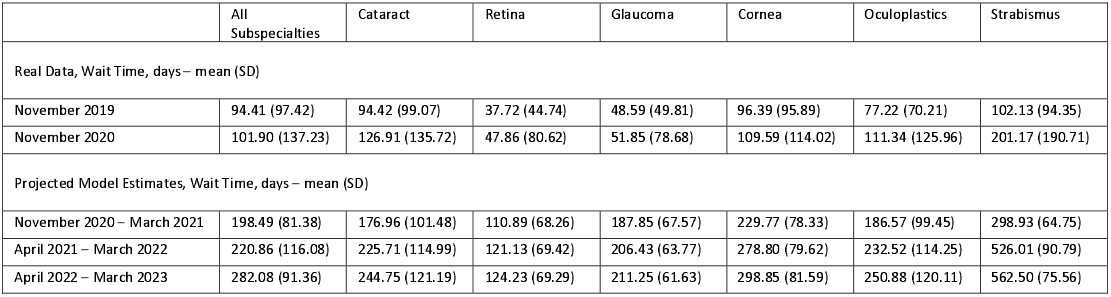
Real data wait times for patients awaiting semi-urgent and non-urgent subspecialty surgery for November 2019 and November 2020. Model-estimated projections for wait times up to 3 years after the pandemic. Scenarios were run 50 times for a total of 240,000 patients.

Regarding backlog clearance, the increase in provincial monthly resources required to clear all surgery types by March 2023 was estimated to be 34% (4,626 additional surgeries per month, Recovery Plan A) if starting in September. Comparatively, recovery to the pracademic waitlist over a shorter period of time until March 2022 would require an increase of 87% (11,838 additional surgeries per month, Recovery Plan B) if starting in September (Figure 2).

## Interpretation

This study aimed to predict the impact of the COVID-19 pandemic on the backlog for ophthalmic surgeries in Ontario, Canada. Following March 15, 2020, the pandemic shutdowns resulted in reduction of surgical volumes for several months and an associated increase in the number of patients awaiting surgery. Similarly, drastic reductions by 77-90% of the usual volume of surgeries performed have been noted at other tertiary ophthalmic surgical centres in North America^23^ and Europe.^25^ Despite the gradual recovery in surgical activity, without any substantial increases in resources to support the backlog of surgical cases, incoming new urgent cases will lead to further delays in surgeries for semi-urgent and non-urgent cases. In addition to modelling the projected backlog as a result of the pandemic, we forecast potential recovery planning scenarios. Our model estimates ophthalmology resource use and availability based on priority level for each subspecialty surgery, while taking into account the increasing urgency over time due to the deterioration expected with delayed access to care. The validity of our study is strengthened by incorporating historical data from the Wait Time Information System database and the local health care resources estimates of potential capacity as well as the use of evidence-based guidelines on established prioritization for subspecialty surgery.

One of the important considerations of delay to surgery is the deterioration in vision outcomes in patients while awaiting surgery. For the patients on the wait list at the time of the pandemic, on average there was a delay of 197 days until surgery. Although with the increase in the number of surgical procedures performed during the post-pandemic recovery phase, there will be a progressive plateau of the number of cases awaiting surgery on the wait list, it is important to note that non-urgent and semi-urgent cases exceeding the acceptable wait times for surgery may progressively deteriorate and become urgent. For example, the cases awaiting sub-specialty surgery for retina had an average of 48 days of wait time immediately following the pandemic, with a substantial increase to 121 and 124 days at 2 and 3 years, respectively following the pandemic. These progressive increases in wait time for non-urgent and semi-urgent cases such as elective epiretinal membrane peel or macular hole repair may lead to consequent vision loss and/or more challenging surgical repairs.^25,26^ Within the realm of other common procedures, recent studies have also reported an average delay of 5.34 weeks as a result of COVID-19 lockdowns for patients requiring intravitreal injections.^27^ The reported implications of this delay were more profound vision loss in patients with diabetic macular edema, proliferative diabetic retinopathy and retinal vein occlusion.^27^

Amongst all subspecialty ophthalmology surgery, cataract is the highest volume of cases (12,697) added to the wait list each month. Aside from the effects of the pandemic, there is an increasing need for higher volumes of cataract surgeries performed with the growth and aging of the population. Hatch et al projected a minimum of 128% increase in surgical volumes or approximately a 4.3 million additional cataract surgeries required per year for 2036 in North America.^29^ The delay in semi-urgent and non-urgent procedures such as cataract surgery, not only have implications on quality of life,^29^ but have also been shown to be associated with increased falls in patients awaiting surgery.^30^ The psychological and physical trauma associated with increased wait times have also been noted by studies on other surgical subspecialty care.^31^ With the consideration of implications of delay in surgery and economic benefits, several have advocated for bilateral surgeries^32^ and combined procedures such as phacovitrectomy.^33^

In addition to the delay to surgery while on the wait list, there is an inherent delay in presentation to specialized surgical care.^14,35,36^ These delays are shown to be further exacerbated by the COVID-19 pandemic and patient hesitation to seeking care in the ophthalmology setting.^7,8^ In this model, the number of adult patients awaiting surgical intervention for strabismus grew notably by 340% within two years. Adult strabismus surgery is rarely considered to be of urgent priority compared to other ophthalmic subspecialty surgery. Nonetheless, this patient population already experiences delays in presentation with approximately 20 years from time of onset to seeking surgical intervention.^37^ As such further delays for strabismus surgery as a result of the COVID-19-induced surgical backlog will result in social, psychological, and economic burden for patients.^37^

With the rapidly evolving nature of the pandemic and unpredictable impact on the healthcare system, it is paramount that decision-makers and government representatives continue using models as tools for evidence generation in support of the policy decision-making processes.^10^ Previous models by our group, have guided decisions regarding the estimation of COVID-19-induced depletion of hospital resources^11^ as well as policies for transmission risk in schools versus community-based settings.^38^ Other models on COVID-19 elective vascular surgical delays have estimated an 8 month recovery period to achieve a steady state in the number of patients awaiting surgery.^39^ Additionally, we have estimated incremental growth in waitlist for all cardiac procedures during the COVID-19 pandemic and the implications for the provision of cardiovascular care.^40^ Wang et al, estimated an additional 719 hours of weekly operating room time to clear the backlog created as a result of the pandemic for surgical specialties such as ophthalmology, gynecology, general surgery, orthopedics and urology.^1^ This model, however, did not investigate ophthalmology in detail and underscored the impact of continued delay on vision outcomes of patients and consideration of interfacility transfer of cases to hospitals with available capacity.^39^

The projections of required resources for the future are essential for introducing policies such as implementation of operating room hours on weekends and extended hours on weekdays. Our projected model estimates suggest that a planned increase in provincial monthly resources of at least 34% will be required to return to the pre-pandemic surgical backlog by March 2023, while also caring for the normal flux of surgical patients. To ensure that this ramp-up in resources is achieved, one possible way to increase volume of surgeries perform will include potentially utilizing a typical 8-hour day or a 40-hour week with an additional 2 hours added to 2 working weekdays as well as one full weekend workday for surgical cases. This recovery planning could be dramatically enhanced if hospitals were to share the burden of cases across the province. Other considerations such as the operational directives regarding patient transport, operating room preparation, personnel dressing and environmental sanitization should be established in place to improve efficiency of the surgeries performed.^6,42^

One of the major strengths of the microsimulation model is the use of comprehensive databases that accurately captured the impact of the pandemic on surgical centres. Furthermore, we had access to detailed information on cases from several facilities. Modeling is a validated and useful tool for providing evidence to support policymakers and decision-making throughout a pandemic.^10^ Our validation results provided strong support that the current model estimates are comparable to historical data. The complex dynamic navigation of patients and interaction between resource availability and demand from the population for the future is most accurately estimated using microsimulations such as in this model. This is the first ophthalmology surgery microsimulation model to consider the prioritization of patients based on wait time and take into consideration and impact of delayed surgery on surgical priority based on the unique characteristics of each of the surgical subspecialties.

This model can be used for other scenarios, with new interruptions in ophthalmic surgeries. With the concerns of additional lockdowns in the future, it is important to note the competing resource allocation for various other procedures and surgical specialties.^4,42,43^ It is proposed that long-term approaches such as increased accessibility to anesthesia assistants and operating room nursing staff as well as additional operating room space may ensure ramp-ups in surgical volumes are quickly re-enacted, and that the health care system could rapidly adjust to increasing demands due to the pandemic and beyond.

### Limitations

It is important to note that this model relies on forecasting COVID-19 cases based on historical data, current evidence-based guidelines and assumptions of patterns in surgical practice. Actual data about the number of patients awaiting surgery beyond March 2020 is currently not available due to the reductions in number of patients seeking healthcare during the pandemic. Although we aimed to capture the population growth rates of 1.3-1.4% when estimating the number of patients presenting for each subspecialty surgery based on historical data, we did not take into consideration other factors such as aging population and evolving changes in the healthcare needs over time which would result in larger volumes of cases performed. This model also assumed that all reported historical surgical volumes were appropriately indicated for surgery, which may not always hold true depending on variations in practice patterns.^44^ Although less common, bilateral surgeries may be performed but the model was set up to simulate each patient eye separately. Data for urgent cases was based on tertiary care centres in the Greater Toronto Area, which likely provides a skewed number of emergencies. Lastly, it is expected that predictions over longer time horizons will be progressively less robust in their reliability.

## Conclusion

The findings from this microsimulation model informed by historical provincial data of surgical volumes depicts the projected wait time for surgery and growing wait list for ophthalmology subspecialty surgery following the COVID-19 pandemic. The recovery plans proposed here for increasing the resources required for clearing the backlog will aid institutions in optimizing their response to the evolving ophthalmic surgical needs of the population.

## Data Availability

Data is not available for external access.

## Contributor’s Statement

Conception and Design: T Felfeli, R Ximenes, D Naimark, S El-Defrawy, B Sander

Acquisition of Data: T Felfeli, P Hooper, R Campbell, S El-Defrawy

Data Analysis: T Felfeli, R Ximenes, D Naimark, B Sander

Interpretation of Data: T Felfeli, R Ximenes, S El-Defrawy, B Sander

First draft of the Article: T Felfeli

Critical Revision: T Felfeli, R Ximenes, D Naimark, P Hooper, R Campbell, S El-Defrawy, B Sander

Final Approval of the Version to be Published: T Felfeli, R Ximenes, D Naimark, P Hooper, R Campbell, S El-Defrawy, B Sander

Act as Guarantor of the Work: T Felfeli, S El-Defrawy, B Sander

## Data-Sharing Statement

Portions of the data are available to others and can be accessing by emailing the corresponding authors.

