## Supplemental Figure 1 for "The Ophthalmic Surgical Backlog caused by the COVID-19 Pandemic: A population-based and microsimulation modelling study"

Total number of patients awaiting surgery based on urgency level (Urgent, Priority 1; Semi-Urgent, Priority 2 and 3; Non-Urgent, Priority Level 4). The simulations were run 50 times (variations in projected estimated represented by grey lines) for a total of 240,000 patients.

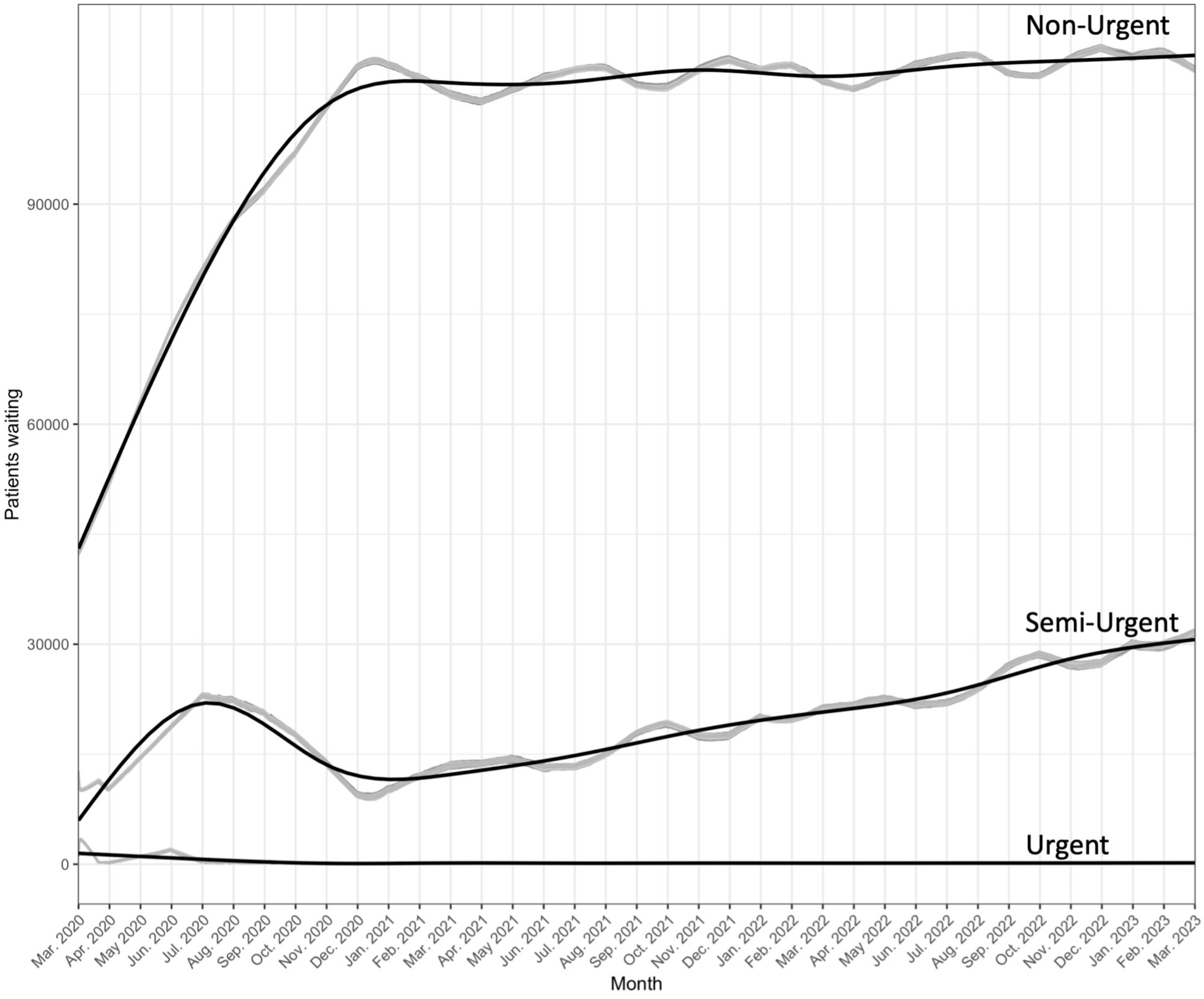
