## Supplemental Figure 2 for "The Ophthalmic Surgical Backlog caused by the COVID-19 Pandemic: A population-based and microsimulation modelling study"

Number of patients awaiting ophthalmic surgery amongst the cohort of patients on the wait list at the start of the pandemic on March 15, 2020. The simulations were run 50 times (variations in projected estimated represented by grey lines) for a total of 240,000 patients.

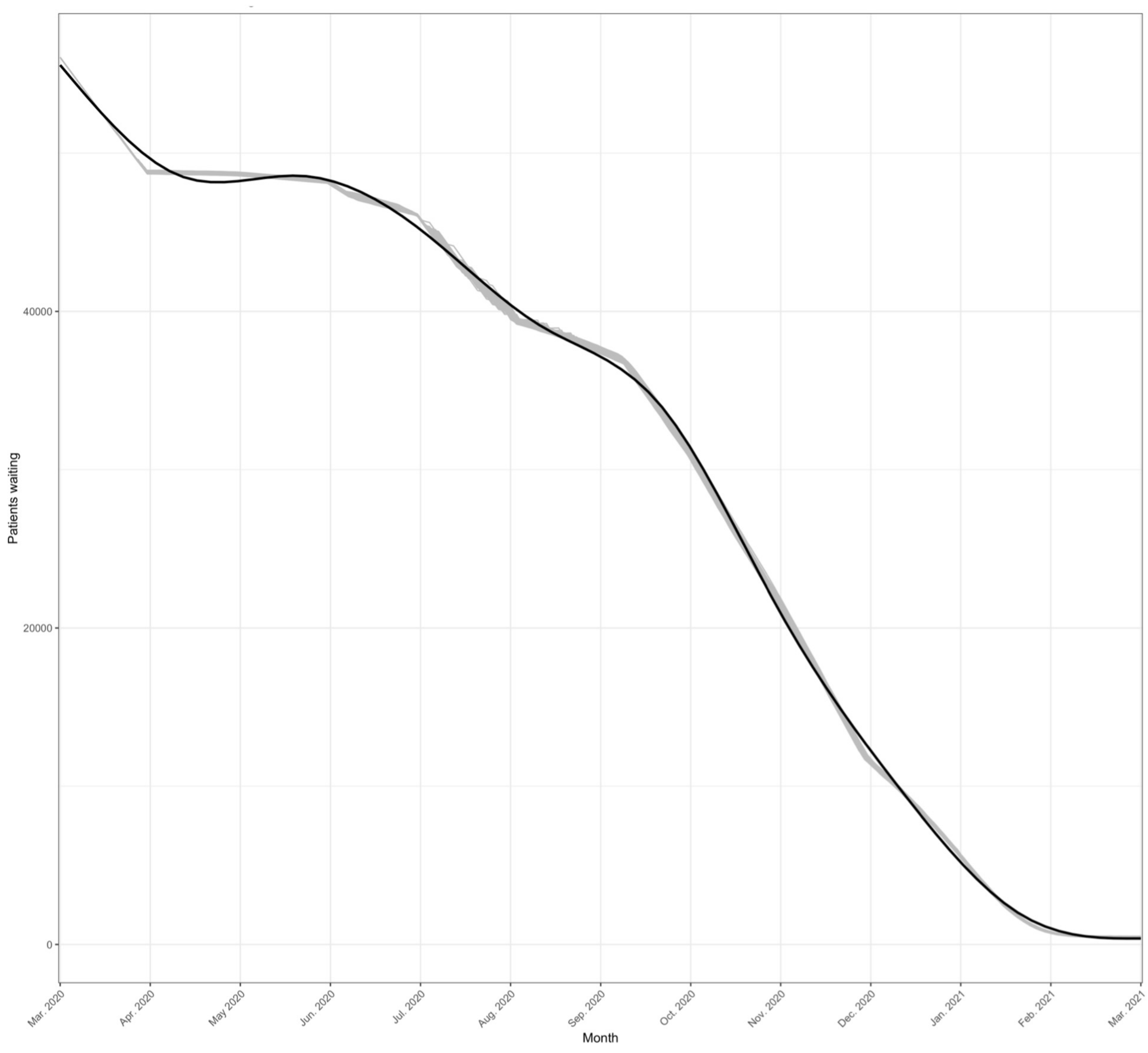
