## Supplemental Table 1 for "The Ophthalmic Surgical Backlog caused by the COVID-19 Pandemic: A population-based and microsimulation modelling study"

**Supplemental Table 1.** Provincial definitions of priority levels for subspecialty surgery types.

| **Priority** | **Description** | | | |
| --- | --- | --- | --- | --- |
| **1** | - Immediate – emergency surgery required   Criteria for urgent cataract cases: | | | |
|  | **Cataract**  Vision impairs activities of daily living and ability to live independently  Vision reduction leading to increased risk of accidental harm  Unable to continue driving  Unable to see faces  Unable to read price tags    Prioritize based on:   1. Whether the patient is monocular 2. Extent of impediment to activities of daily living and living independently 3. Extent of risk of accidents | **Cornea**  Painful bullous keratopathy  Advanced corneal edema with vision below 20/80  Bilateral corneal disease reducing vision and limiting activities of daily living.  Impending perforation/perforation    Prioritize based on:   1. Whether the patient is monocular 2. Severity of pain 3. Extent of improved outcome doing surgery now 4. Risk to the eye if surgery not done now | **Glaucoma**  2 of the following 3:  Significant risk of disease progression with further postponement of surgery  Advanced glaucoma with IOP above target with evidence of progression  IOP markedly elevated on MMT (at any stage of glaucoma)    Prioritize based on:   1. Whether the patient is monocular 2. Severity of field loss/progression 3. Severity of disc changes 4. Level of IOP elevation over target | **Retina**  Surgery now will improve final visual outcome, and /or reduce the risk of endophthalmitis and/or reduce the risk of ocular inflammation  Prioritize based on:   1. Whether the patient is monocular 2. Extent of improved outcome doing surgery now 3. Risk to the eye if surgery not done now |
| **2** | - Moderate probability of disease progression. Low probability of disease occurrence or progression impacting morbidity or mortality. | | | |
| **3** | - All patients who do not meet the criteria of Priority 2 or Priority 4. | | | |
| **4** | - Minimal risk of disease progression impacting morbidity/mortality. | | | |

Source: The Vision Task Force Committee. Quality-Based Procedures Clinical Handbook for Cataract Day Surgery. Ministry of Health and Long-Term Care. http://www.health.gov.on.ca/en/pro/programs/ecfa/docs/qbp_cataract.pdf. Published 2018. Accessed December 28, 2020.
