## Supplemental Table 2 for "The Ophthalmic Surgical Backlog caused by the COVID-19 Pandemic: A population-based and microsimulation modelling study"

**Supplemental Table 2.** Key model input data parameters. Data sources: Facility Level Data; Wait Times Information System, Ontario Health (Cancer Care Ontario); Expert Opinion; Published Guidelines; Literature

| Variables | **Cataract** | **Retina** | **Glaucoma** | **Cornea** | **Oculoplastics** | **Strabismus** |
| --- | --- | --- | --- | --- | --- | --- |
| Prob. of Urgency |  |  |  |  |  |  |
| Urgent | 0.01 | 0.15 | 0.04 | 0.02 | 0.01 | 0 |
| Semi-urgent | 0.24 | 0.12 | 0.04 | 0.05 | 0.2 | 0.01 |
| Non-urgent | 0.75 | 0.73 | 0.92 | 0.93 | 0.79 | 0.99 |
| Prob. of Anatomical Success |  |  |  |  |  |  |
| Semi and Non-urgent |  |  |  |  |  |  |
| Primary Surgery | 0.98 | 0.85 | 0.8 | 0.95 | 0.9 | 0.9 |
| Second Surgery | 0.9 | 0.9 | 0.8 | 0.9 | 0.8 | 0.9 |
| Third Surgery | 0.8 | 0.6 | 0.8 | 0.8 | 0.9 | 0.9 |
| Urgent | 0.85 | 0.85 | 0.8 | 0.85 | 0.85 | 0.85 |
| Prob. Two-step Surgery | 0.03 | 0.03 | 0.03 | 0.03 | 0.03 | 0.03 |
| Target Wait Times* |  |  |  |  |  |  |
| Urgent | 3 | 3 | 3 | 3 | 30 | 183 |
| Semi-urgent | 84 | 14 | 30 | 84 | 60 | 365 |
| Non-urgent | 182 | 90 | 120 | 182 | 180 | 370 |
| Time to Deterioration** |  |  |  |  |  |  |
| Urgent | 6 | 6 | 6 | 6 | 60 | 366 |
| Semi-urgent | 168 | 28 | 60 | 168 | 120 | 730 |
| Non-urgent | 364 | 180 | 240 | 364 | 360 | 740 |

*Sources:

1. The Vision Task Force Committee. Quality-Based Procedures Clinical Handbook for Cataract Day Surgery. Ministry of Health and Long-Term Care. http://www.health.gov.on.ca/en/pro/programs/ecfa/docs/qbp_cataract.pdf. Published 2018. Accessed December 28, 2020.

2. The Vision Task Force Committee. Quality-Based Procedures Clinical Handbook for Integrated Retinal Care. Ministry of Health and Long-Term Care.

3. The Vision Task Force Committee. Quality-Based Procedures Clinical Handbook for Integrated Corneal Transplant Care. Ministry of Health and Long-Term Care. http://www.health.gov.on.ca/en/pro/programs/ecfa/docs/hb_corneal.pdf. Published 2017. Accessed December 28, 2020.

**Cases are prioritized in a higher level of urgency once the maximum ‘Time to Deterioration’ in their initial priority level is reached.
